# From Uncertain to Actionable: Significant Reduction in Variants of Uncertain Significance in Hereditary Germline Testing via Multi-Institutional Real-World Evidence

**DOI:** 10.1101/2025.08.12.25333547

**Authors:** Kelly M. Schiabor Barrett, Matthew J. Ferber, Sophie Candille, Ildiko Thibodeau, Daniela Iacoboni, Amy C. Sturm, Chad R. Haldeman-Englert, Steven F. Powell, Douglas Stoller, Christopher N. Chapman, C. Anwar A. Chahal, Daniel P. Judge, Douglas A. Olson, Joseph J. Grzymski, Nicole L. Washington, Catherine Hajek, William Lee, Alexandre Bolze, James Lu, Elizabeth T. Cirulli

**Affiliations:** Helix, San Mateo, CA; The Ohio State University Wexner Medical Center, Columbus, Ohio, USA; Cone Health Precision Health, Greensboro, North Carolina, USA; Sanford Health, Sioux Falls, South Dakota, USA; University of Nebraska Medical Center, Omaha, Nebraska, USA; Department of Pathology & Laboratory Medicine, St. Luke’s University Health Network, Bethlehem, Pennsylvania, USA; Center for Inherited Cardiovascular Diseases, WellSpan Health, York, Pennsylvania, USA; Department of Cardiovascular Medicine, Mayo Clinic, Rochester, Minnesota, USA; Barts Heart Centre, West Smithfield, London, UK; William Harvey Research Institute, Queen Mary University of London, Charterhouse Square, London, UK; Division of Cardiology, Medical University of South Carolina, Charleston, South Carolina, USA; HealthPartners, Minneapolis, Minnesota, USA; University of Nevada, School of Medicine, Reno, Nevada, USA; Renown Health, Reno, Nevada, USA

## Abstract

The clinical utility of genomic testing is constrained by variants of uncertain significance (VUS), which complicate diagnostic interpretation and patient management. The ACMG/AMP PS4 criterion, “prevalence in affected individuals statistically increased compared to controls,” offers strong evidence for pathogenicity but is often challenging to apply due to the limited availability of robust, matched case-control genomic and phenotypic data. Further, there are currently no options available to score evidence from case-control studies towards benignity.

We propose and validate a new code, RWE (real world evidence), by integrating de-identified, longitudinal clinical data with variant carriers and non-carriers identified from exome or genome sequence data across three large-scale clinicogenomic datasets: the Helix Research Network (HRN) dataset, UK Biobank (UKB) and All of US (AoU). Phenotypes for established gene-level disease associations were compiled from the longitudinal medical records of the individuals, enabling rigorous variant-specific case-control analyses from population data. This RWE approach was systematically applied to all variants, including previously identified VUS in clinically relevant genes, powering our VUS Early Surveillance Platform.

Across 20 hereditary cancer and cardiovascular genes, the application of RWE provided sufficient evidence to reclassify a VUS in 32% of VUS carriers–99.7% to B/LB and 0.3% to P/LP–directly resolving their ambiguous status. This reclassification rate varied by gene, ranging from 0.7% for *BRCA2* up to 50% for *LDLR*.

The systematic integration of Real-World Evidence from large-scale clinicogenomic datasets into the ACMG/AMP scoring rubric through our newly developed and statistically robust RWE category is a significant improvement to variant interpretation that is projected to resolve over 50% of VUS carriers once longitudinal clinico-genomic databases are available for ~3M individuals. This approach markedly reduces the burden of Variants of Uncertain Significance, provides more definitive diagnoses for a substantial proportion of previously unresolved cases, and ultimately increases the clinical utility and adoption of genomic testing, representing a critical advancement for precision medicine.

## Introduction

The advent of next-generation sequencing (NGS) has revolutionized molecular diagnostics. Gene panel testing, clinical whole exome sequencing (WES), and whole-genome sequencing (WGS) are now integral to identifying disease-causing variants for Mendelian disorders, guiding therapies and treatment choices as well as informing long-term health outlooks for families. While diagnostic testing can clarify diagnosis and prognosis for patients, classifying variants as variants of uncertain significance (VUS) can often create confusion for patient management. This classification often results in over-treatment and delays in appropriate medical management, and can cause considerable anxiety for patients and their families, motivating Helix to establish the VUS Early Surveillance Platform ^1,2^.

The VUS classification is one of five classifications utilized in the prevailing variant interpretation framework, jointly developed by the American College of Medical Genetics and Genomics (ACMG) and the Association for Molecular Pathology (AMP)^3^. This framework was developed to encourage standardization. It works by aggregating scores from different evidence types (e.g., population data, computational predictions, functional data, case reports, segregation data, and de novo occurrence). The scores sum to either a Pathogenic (P), Likely Pathogenic (LP), VUS, Likely Benign (LB), or Benign (B) interpretation. Recent research re-evaluating historic VUS interpretations have shown at least 80% of variants that fall into this category ultimately reclassify to benign, suggesting over usage of this category ^4^. Ideally, laboratories would avoid usage of the VUS category when possible. Certainty in either direction provides clarity to clinicians on how best to proceed with patient management.

One major driver of classification updates has been the open-source availability of variant frequency data from population-based studies (captured in the ACMG/AMP rubric via PM2, BS2, BS1, and BA1 categories). The use of these data follow the basic logic that if a variant is more common than the disease itself, that variant is likely not individually causative for the condition in question. Variant frequency data alone, however, only provides contributory evidence. Most monogenic conditions, particularly in adults, manifest over time. Variant information alone, without other phenotypic information, may not appropriately account for the natural history of disease, where symptomatic disease detection may be clinical observation of a long latent condition driven by genetic risk. Clinicogenomic data, which systematically combines variant data with longitudinal clinical information, has the potential to take this type of assessment dramatically further. Longitudinal phenotypes, including the putative age of first presentation, can be utilized to better understand the natural course of disease for any particular variant, pathogenic and benign.

Current ACMG/AMP guidelines attempt to include these types of data under the PS4 criterion, “the prevalence of the variant in affected individuals is significantly increased compared with the prevalence in controls”^2^. However, applying the PS4 criterion rigorously is often challenging, with guidance for usage varying widely by condition. At its core, this criterion requires access to well-phenotyped case cohorts and appropriately matched control populations of sufficient size and depth to achieve statistical power, analyzed with statistical methods appropriate for genetic data. This is especially challenging for individually rare variants that nonetheless contribute to a variety of disease conditions and incidence rates. Such datasets, particularly those linking genomic information with longitudinal clinical phenotypes across populations, are not readily available to diagnostic laboratories, and as a result, this type of evidence is often applied via compilations of case series and observational studies instead of through more robust association analyses^5^.

Longitudinal Real-World Data (RWD), derived from sources such as electronic health records (EHRs), insurance claims, and patient registries, offer a transformative opportunity to generate Real-World Evidence (RWE) for large, diverse patient populations regardless of disease status ^6^. The systematic integration of RWD with genomic data holds the potential to establish robust case-control cohorts for evaluation of specific gene-disease pairs, thereby enabling a more thorough application of criteria like PS4. Further, establishing population cohorts allows the extension of this approach to score variants towards benignity–PS4 is only scoped for evidence of pathogenicity.

This type of data does not have a clear “home” in the current ACMG/AMP rubric. Our study describes the development and application of a new category for this unique data source, which we call real world evidence (RWE) and utilize in our VUS Early Surveillance Platform.

Leveraging three large, de-identified real world clinicogenomic datasets from the general population, we identify statistically significant deviations in variant prevalence among clinically-defined affected individuals compared to controls, controlling for important covariates including age, thereby providing crucial new evidence to reclassify VUS towards both Benignity or Pathogenicity. We demonstrate the utility of this RWE category through its application to variants in clinically relevant genes across both cardiovascular and cancer applications, leading to a dramatic reduction in VUS rates and an increase in the clinical utility of genomic testing.

## Methods

### Data Source and Study Populations

Helix Research Network™ (HRN) data came from 227k participants who were recruited from 9 North American health systems as of Feb 2025: Ohio State Genomic Health (OSU Wexner Medical Center, Ohio), DNA Answers (St. Luke’s University Health Network, Pennsylvania), GeneConnect (Cone Health, North Carolina), the Genetic Insights Project (Nebraska Medicine), the Healthy Nevada Project (Renown Health, Nevada), ImagineYou (Sanford Health, Upper Midwest), In Our DNA SC (Medical University of South Carolina), myGenetics (HealthPartners, Minnesota), and The Gene Health Project (WellSpan Health, Pennsylvania)^7^. HRN samples were sequenced and analyzed at Helix using the Exome+^®^ assay as previously described^7,8^.

Data were processed using a custom version of Sentieon and aligned to GRCh38, with variant calling and phasing algorithms following GATK best practices. The Helix cohort studies were reviewed and approved by their applicable Institutional Review Boards, and all participants gave their informed, written consent prior to participation.

From the UKBiobank (UKB), we used population-level exome OQFE pVCFs (field 23157), as previously described ^9^. The UKB study was approved by the North West Multicenter Research Ethics Committee, UK. All of Us (AoU) whole genome sequence data were restricted to the exonic regions using the v7 plink bed resource ^10^. The All of Us study is approved by the All of Us IRB.

### Phenotype Definition

For HRN, International Classification of Diseases, Ninth and Tenth Revision ICD codes and associated dates (ICD-9 and ICD-10-CM) were collected from available diagnosis tables (from problem lists, medical histories, admissions data, surgical case data, account data, claims, and invoices). For the UKB, ICD codes and associated dates (both ICD-9 and ICD-10) were collected from inpatient data (category 2000), cancer register (category 100092) and the first occurrences (category 1712). For All of Us, the phenotypes from the All by All phecode, lab measurements, and drug resources were used ^11^.

Phecodes were used for breast (CA_105) and ovarian (CA_106.3) cancer, and the analysis was restricted to females for these phenotypes^12^. Phecodes were also used for colon (CA_101.41) and endometrial (CA_106.21) cancer. For familial hypercholesterolemia, the median of all available LDL measurements per individual were used, normalized using rank-based inverse normal transformation. Statin use was included as a covariate when analyzing LDL levels.

### Genetic Data

Variant annotation was performed with VEP 104 ^13^. Coding regions were defined according to Gencode version GENCODE 33, and the MANE transcript was used to determine variant consequence ^14,15^. Genotype processing for HRN data was performed in Hail 0.2.115-10932c754edb and plink ^16,17^. Variants included were those that were exonic–including synonymous variants but not UTR–and those at splice acceptor or donor sites. Missense variants were considered damaging at a REVEL score>0.75 or if they were in-frame insertions or deletions. LoF variants included stop gain, start gain, stop loss, and frameshift indels.

Variant interpretation for small indels and single nucleotide variants was completed using an automated, two-step approach. First, a variant was considered either Pathogenic (P/LP) or Benign (B/LB) If it had a known and well-established clinical Pathogenic or Benign interpretation (i.e., no variant of uncertain significance (VUS) or conflicting interpretations present in ClinVar across high volume laboratories, using search strings [‘ClinGen’, ‘Quest’, ‘Sema4’, ‘Natera’, ‘Invitae’, ‘All of Us’, ‘Baylor’, ‘GeneDx’, ‘Ambry’, ‘LapCorp’, ‘Color’, ‘Myriad’, ‘Brigham’] and/or a P/LP or B/LB interpretation by the relevant variant curation expert panel (VCEP); InSiGHT Hereditary Colorectal Cancer/Polyposis VCEP, Evidence-based Network for the Interpretation of Germline Mutation Alleles (ENIGMA) *BRCA1* and *BRCA2* VCEP, and the Familial Hypercholesterolemia VCEP. 9,621 variants had well-established clinical interpretations. Additionally, for all variants, ACMG-AMP variant interpretations were completed programmatically (automated ACMG) following the gene-specific scoring recommendations from each respective VCEP (when available) or generalized scoring guidance^3^. As input, MANE transcripts were used to define each gene and interpretations leveraged the following tools: VEP-104^13^, GnomADv3^18^, REVEL^19^, SpliceAI^20^, ClinVar database (accessed: 03/10/2025), for *MSH2* functional scores from MAVE (urn:mavedb:00000050-a)^21^, as well as all data provided by each respective VCEP (e.g. domains, functional scores and weights for particular variants, NMD boundaries). In total, the following criteria were assessed using this data: PVS1, PS1, PS3_Supporting, PM1, PM2, PM5, PP3, BA1, BS1, BP4, and BP7. Data from individual case studies as well as patient-specific information such as presenting symptoms or family history were not considered for these interpretations. Negative and positive points, weighted for supporting (1 point), moderate (2 points), strong (4 points) and very strong (8 points), were used to accumulate evidence for both pathogenicity and benignity across these categories^22^. In total, variants with <=−2 points were considered B/LB and variants with >=6 points were considered pathogenic as detailed in prior work ^23^. Data from individual case studies as well as patient-specific information such as presenting symptoms or family history were not considered for these interpretations. For variants with a well-established Clinical interpretation, if there was a discrepancy with the automated ACMG interpretation, the Clinical interpretation was chosen. All automated ACMG scores as well as resulting interpretations are available in **Table S2** (counts and odds ratios filled as −9 when <20 in a category).

### Statistical Analysis and Automated PS4_RWE Evaluation

As previously described, a machine learning whole genome regression model was built from a representative set of 184,445 coding and noncoding LD-pruned, high-quality common variants to account for relatedness and population stratification and ancestry^7,24^. Covariates included 10 ancestry-specific principal components, age and sex, as well as statin use for the LDL analysis. Analyses were performed separately in each ancestry in each cohort and then meta analysis across ancestries and across the three cohorts was performed using METAL^25^.

Strength levels for the RWE criterion were assigned based on the calculated odds ratio and the number of individuals carrying the variant, following an evidence framework adapted from ClinGen recommendations. Effect cutoffs were based on VCEP for HBOC and properties of known pathogenic variants for LS and FH. Additionally, minimum “n” carrier cutoffs were established to reduce false positives. For FH, this was a minimum of 5 (strong) or 3 (moderate) phenotyped individuals. For benign breast/ovarian cancer and LS variants, carrier cutoffs were chosen to expect to see at least 5 (strong) or 2 (moderate) case carriers if the variant’s effect size is equal to the effect size for known pathogenic variants. For HBOC this was an OR of 4 and prevalence of 7.6%; for breast cancer an OR of 2 and prevalence of 6.9%, and for LS an OR of 6 and prevalence of 1.1%. Thresholds were set as in **Table S1**. Likelihood ratios (LRs) were calculated as the ratio between the sensitivity and 1-specificity for known pathogenic variants in each category (**Table 2**).

**Table 2.**
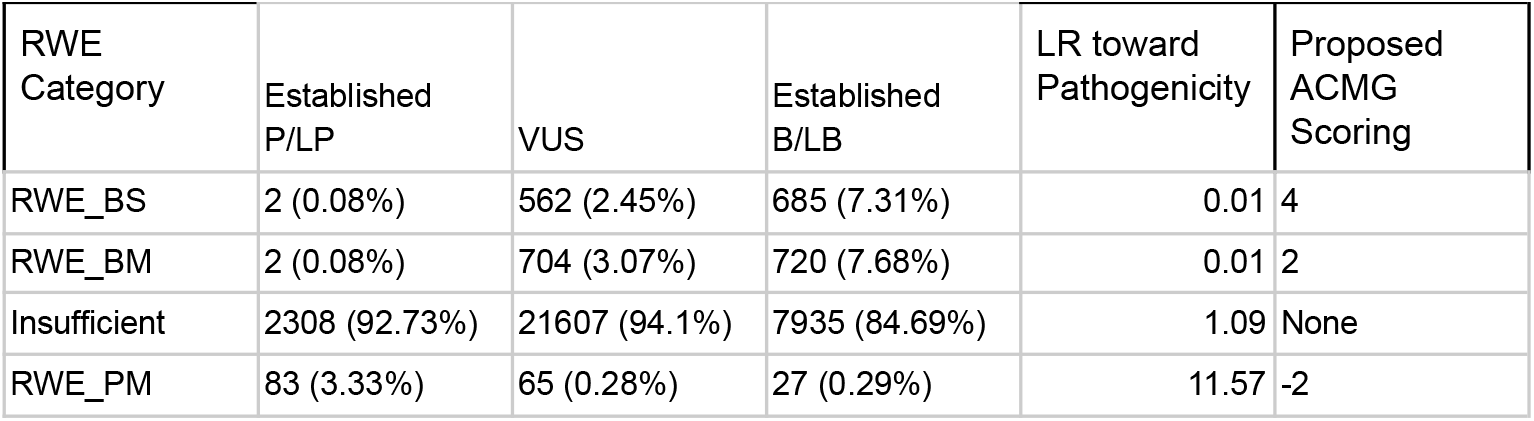
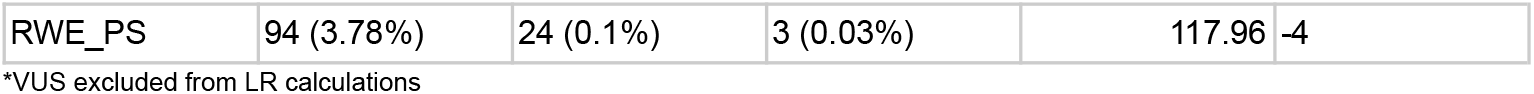
Alignment between RWE and established Pathogenic and Benign calls.

| RWE Category | Established P/LP | VUS | Established B/LB | LR toward Pathogenicity | Proposed ACMG Scoring |
| --- | --- | --- | --- | --- | --- |
| RWE_BS | 2 (0.08%) | 562 (2.45%) | 685 (7.31%) | 0.01 | 4 |
| RWE_BM | 2 (0.08%) | 704 (3.07%) | 720 (7.68%) | 0.01 | 2 |
| Insufficient | 2308 (92.73%) | 21607 (94.1%) | 7935 (84.69%) | 1.09 | None |
| RWE_PM | 83 (3.33%) | 65 (0.28%) | 27 (0.29%) | 11.57 | -2 |
| RWE_PS | 94 (3.78%) | 24 (0.1%) | 3 (0.03%) | 117.96 | -4 |

\*VUS excluded from LR calculations

## Results

### Variant properties

Overall, 853,503 individuals from the general population with medical records or blood labs available were included in the analysis. Twenty genes were chosen for analysis based on inclusion in common CDCT1 condition lists or hereditary breast cancer lists. All coding and synonymous variants with MAF<1% were considered for the analysis. This included 34,821 variants, with a median of 1,244 variants analyzed per gene.

Variants were analyzed against appropriate gene-level phenotypes across the HRN, UKB, and AoU and classified according to their statistical evidence (**Table 1**). Of the 34,821 variants, 4.1% had strong statistical evidence toward Benign (RWE_BS), 3.6% moderate statistical evidence toward Benign (RWE_BM), 0.5% moderate statistical evidence toward Pathogenic (RWE_PM), and 0.35% had strong statistical evidence towards Pathogenic (RWE_PS), with the remaining having insufficient evidence for any of these categories.

Overall, variants more often had evidence toward Benignity when they were synonymous (8.7%) or non-damaging missense (8.3%), and very few variants in these categories had evidence toward Pathogenicity (0.3% and 0.4%, respectively; **Figure 1**). Of damaging missense variants, 3.7% had evidence toward Benignity, and 2.4% had evidence toward Pathogenicity. For splice acceptor and donor variants, 3.5% had evidence toward Benignity and 2.6% towards Pathogenicity. For other LoF variants (stop gain, loss, or frameshift), 4.7% had evidence toward Benignity and 4.6% toward Pathogenicity. Nearly all the LoF variants with statistical evidence toward Benignity were in *APOB* and *PCSK9*, consistent with these being the only genes in the analysis where LoF does not confer disease.

**Figure 1.**
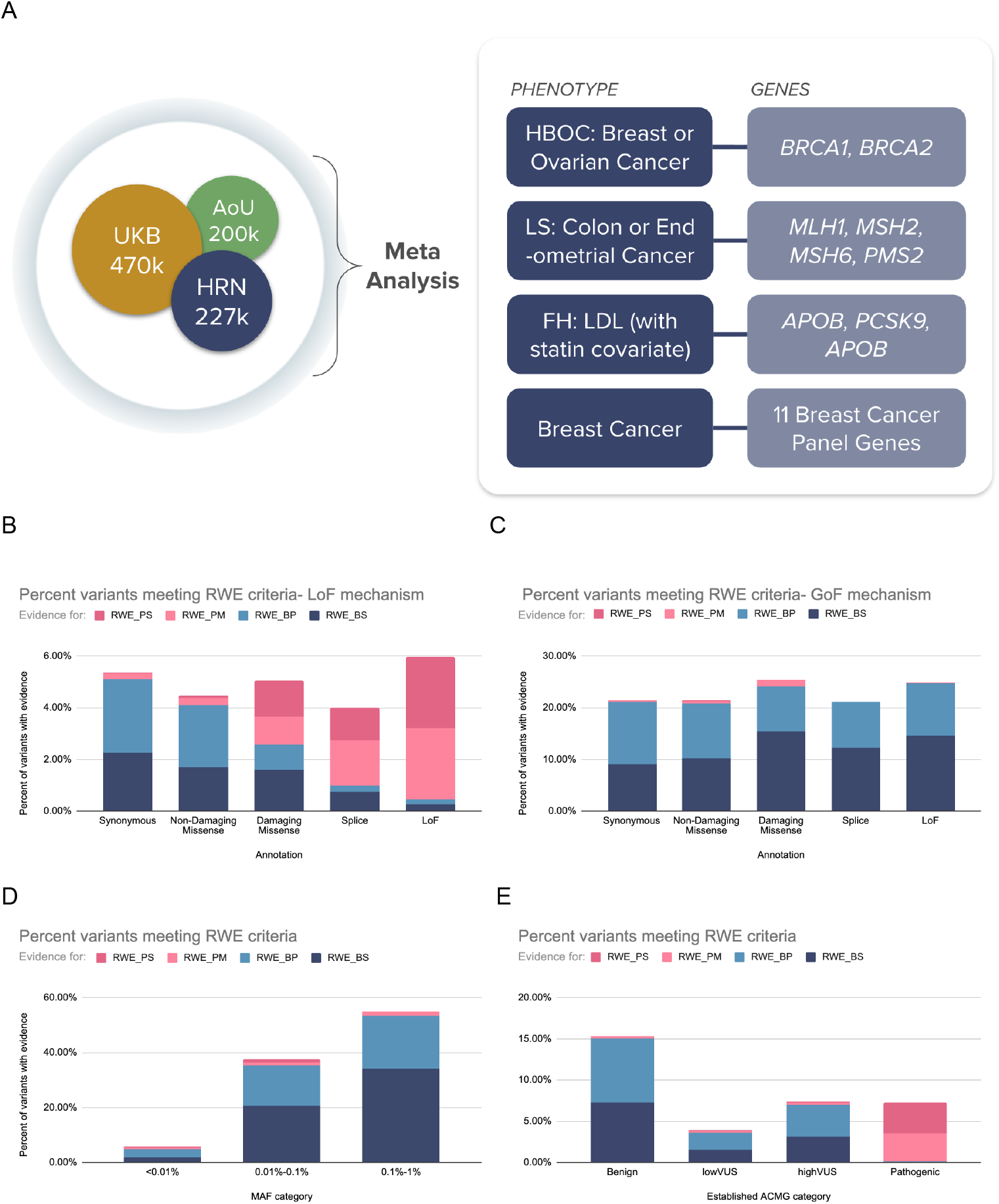
RWE study setup and percent of variants meeting criteria to apply RWE. A) Overview of study sample, genes and phenotypes analyzed. Shown in B-E are the percent of variants with RWE toward each category in different subsets of the data. RWE_PS = Strong Pathogenic Real World Evidence; RWE_PM = Moderate Pathogenic RWE; RWE_BM = Moderate Benign RWE; RWE_BS = Strong Benign RWE. B) and C) split by variant annotation. Damaging missense is those with REVEL>0.75 or in-frame insertions and deletions; splice variants are splice acceptor or splice donor; LoF are stop gain, start gain, start loss, and frameshift. B) Genes with LoF as the disease mechanism. C) Genes with GoF as the disease mechanism (*APOB, PCSK9*). D) Split by MAF. E) Split by established pathogenicity category from ACMG criteria without RWE. lowVUS is −1 or 0 points, highVUS is 1-5 points. Pathogenic indicates Pathogenic or Likely Pathogenic; Benign indicates Benign or Likely Benign.

The amount of evidence toward Benignity and Pathogenicity was strongly influenced by the MAF of the variant and thus the number of people who were informative for the analysis. For example, 55% of variants with MAF 0.1-1% met criteria to apply RWE, while this was only true for 6% of variants with MAF<0.01%. As expected, increasingly lower MAF thresholds become powered for analysis as sample sizes grow.

### Comparison with established Clinical variant interpretations and ACMG Classifications

Of the 34,821 variants analyzed here, 9,975 had well-established clinical interpretations and 24,846 required automated ACMG/AMP scoring to establish an interpretation (see Methods). Without incorporating RWE, 2,489 can be classified as Pathogenic or Likely Pathogenic, 9,370 as Benign or Likely Benign, and 22,962 as VUS. Of the established Pathogenic variants, the statistical evidence from RWE confirms Pathogenicity for 7.1%, supports Benignity for 0.2%, and has insufficient evidence in the other 93% (**Table 2**). Of the established Benign variants, 15.0% had RWE evidence in support of Benignity, 0.3% for Pathogenicity, and 85% without clear evidence either way (**Table 2**). Of the VUS group, 5.5% had RWE evidence in support of Benignity, 0.4% in support of Pathogenicity, and 94% without sufficient evidence. The similarly low percentage of VUS and Benign variants with evidence toward pathogenicity corroborates accumulating evidence indicating that the VUS group is over-indexed for what are likely benign variants. These trends held splitting VUS into higher and lower scoring variants as well as those classified by ClinVar vs. automated ACMG/AMP scoring (**Figure 1**; **Figure S1**).

To calibrate our results to ACMG scoring, we next calculated likelihood ratios (LRs) for each statistical category, using either the established pathogenic and benign clinical calls or all variants with a pathogenic or benign interpretation as a source of truth (see Methods; **Table 2**; **Table S3**). Across both data sets, we find the LRs for our high statistical support categories in both directions match expectations for strong level of evidence: LR of 118 for RWE_PS compared to an expected LR 18.72-350.4 for PS; and LR of 0.01 for RWE_BS compared to an expected LR 0.000285-0.053 for BS^22,26^. Our variants with medium statistical support meet or exceed expectations for moderate evidence: LR of 12 for RWE_PM compared to an expected LR 4.33-18.72 for PM; and LR of 0.01 for RWE_BM compared to an expected LR 0.053-0.23 for BM. Because of this alignment with existing LR patterns, we suggest these categories to be considered at the same evidence level and as existing ACMG criteria for evidence at this level.

### Potential to reclassify VUS interpretations with RWE

We then investigated what impact this RWE would have on patients if used to inform variant interpretation. Focusing on HRN, we found that 22.8% of participants had a VUS in at least one of the 20 analyzed genes. The percentage of VUS variants meeting RWE criteria varied widely by gene, with well studied genes like *BRCA1* and *BRCA2* having very few VUS meeting RWE criteria (<5%), and other genes like *LDLR, PCSK9*, and *APOB* having a very high percentage with such evidence (>60%; **Figure 2**). In total across all genes, 37.9% of individuals that had a VUS had RWE evidence toward Benignity, and 1.4% had a variant with evidence toward Pathogenicity (**Figure 2**).

**Figure 2.**
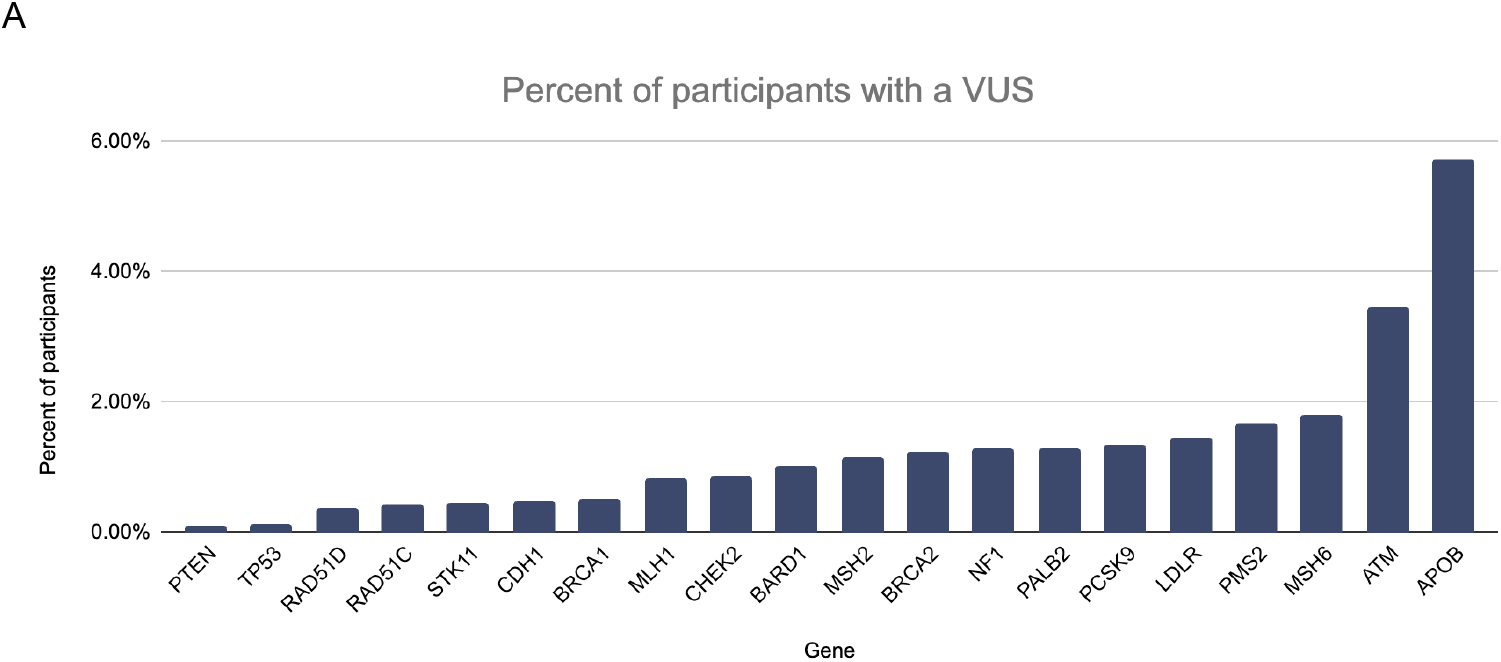
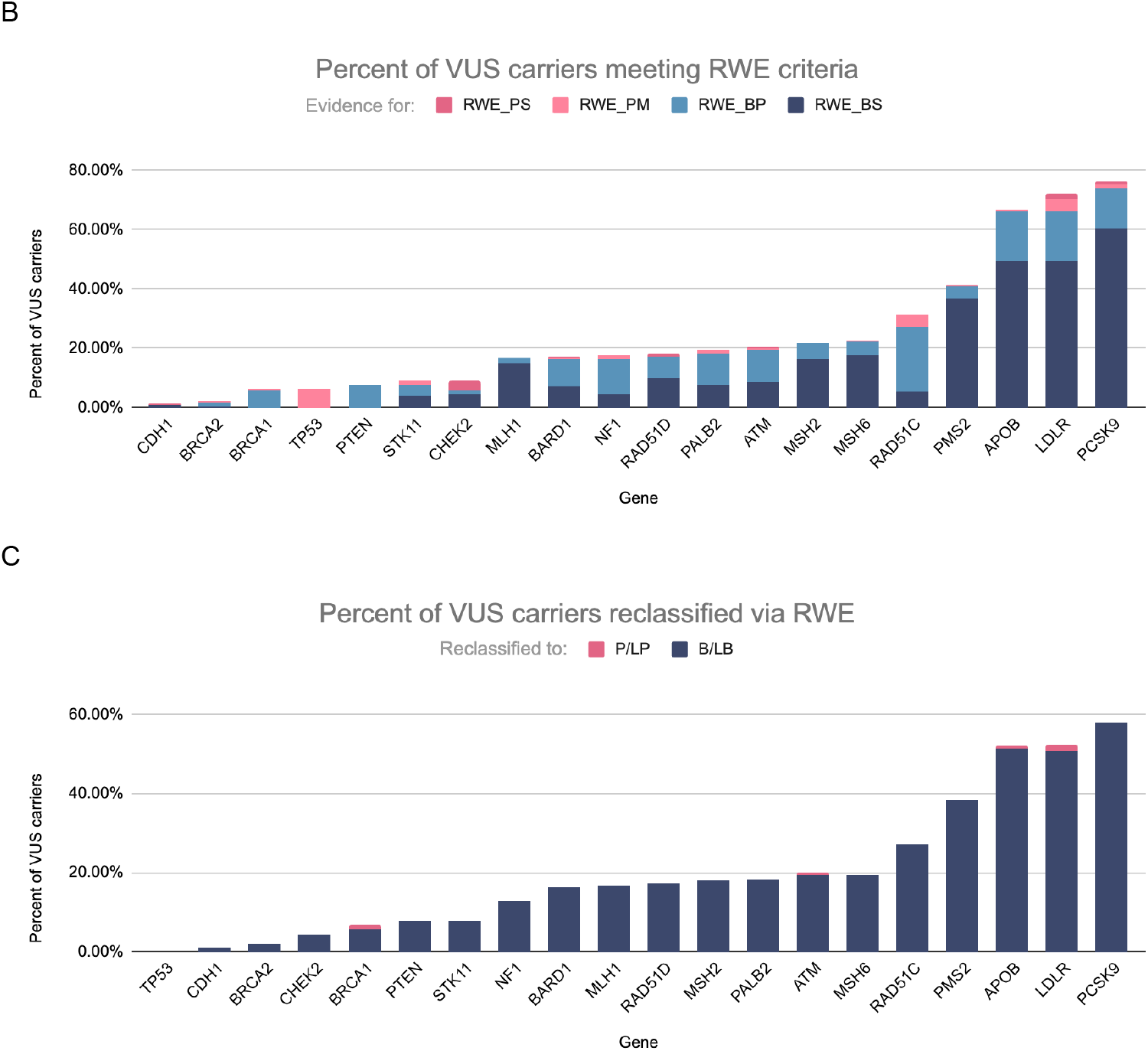
Evidence to reclassify VUS. A) Percent of individuals with an established Pathogenic, Benign, or VUS variant per gene. B) Percent of individuals with a VUS that meets RWE criteria, per gene. C) Percent of individuals with a VUS that gets reclassified when RWE data are included, with strong evidence adding 4 points and moderate adding 2.

Considering the RWE in combination with the points from other ACMG categories, giving 2 points for moderate RWE and 4 for strong RWE, we find that 31.5% of VUS carriers have sufficient evidence from RWE to move into the B/LB (31.4%) or P/LP (0.1%) categories, demonstrating significant potential for this data source. By functional annotation, this reassignment applied to 9.7% (46/476) of individuals with a LoF VUS, 59.3% (33/59) with a splice VUS, 31.9% (16,957/53,084) with a missense VUS, and 30.6% (633/2,066) with a synonymous VUS.

### Projection of evidence generation as RWE datasets grow

The ability to generate statistical evidence for associations with genetic variants is directly related to the number of times the variant is seen in the dataset (**Figure 1**). As datasets grow, a larger number of VUS will have sufficient data to be analyzed via RWE. In the current dataset, 5.9% of VUS had sufficient sample sizes and evidence available for a robust analysis, representing 39% of VUS carriers. Based on the relationship in the present dataset between carrier counts, sample size, and RWE rates, increasing this sample size to 3M individuals is expected to lead to statistical evidence toward reclassification of an additional 2% of VUS, representing an additional 15% of VUS carriers. This would potentially allow for the reclassification of more than 50% of VUS carriers. These gains will continue as datasets grow further, although there is an expectation that there will be an asymptote for the very long tail of novel singletons seen in new patients.

## Discussion

Variant of uncertain significance (VUS), a common variant interpretation, is a very difficult result to manage from a clinical perspective. To advance the integration of genomics into medicine, it’s important that we continue to find ways to limit the number of variants that fall in this category. Large clinicogenomic datasets provide a window into the natural evolution of disease. Even if each record is incomplete, the collective longitudinality of this data is incredibly important in determining if a person with a certain genetic variant is likely to manifest disease and if so, when. In this work, we showcase how real world data of this style can be used in applications such as Helix’s VUS Early Surveillance Platform to build both benign and pathogenic assertions for variants. We propose a new real world evidence (RWE) category for the ACMG/AMP variant interpretation rubric to capture information from this data source.

Many of the benign data categories in the ACMG/AMP rubric relate to variant frequency considerations, leaving limited avenues to establish a benign interpretation for rare variants. We show that RWE can help provide an additional datapoint for both rare and more common variants and has the potential to swing an otherwise uncertain interpretation towards a more definitive benign or pathogenetic result. In particular, examining the natural course of history of VUS in twenty clinically-relevant genes, we find that, as a group, the RWE for VUS variants, even split into higher and lower scoring VUS variant groups, is much more similar in profile to the RWE seen for established Benign variants than the profile of established Pathogenic variants. This result suggests current clinical management should assume a high probability of benign for any reported VUS and highlights the potential to clean up the VUS category.

We believe creating a standalone category for statistical associations from large population datasets is important for several reasons. RWE can be leveraged for all types of variation and will become increasingly relevant and precise for more variants as real-world datasets grow in depth, size, and number (Figure 3). While the study design to analyze these datasets is case-control or quantitative in format, the data source itself is very different from many case-control analyses currently used to inform diagnostic variant interpretation, which often have much less information about the control population used than the cases and may not fully account for population structure. For example, the ability to use RWE for both claims of pathogenicity and benignity is unique to these large population cohorts. Further, this solution leaves the existing category of PS4 available for more diagnostically focused case-driven studies and in particular for rare disease phenotypes that will not be readily captured or easily studied in larger, more unselected datasets.

**Figure 3.**
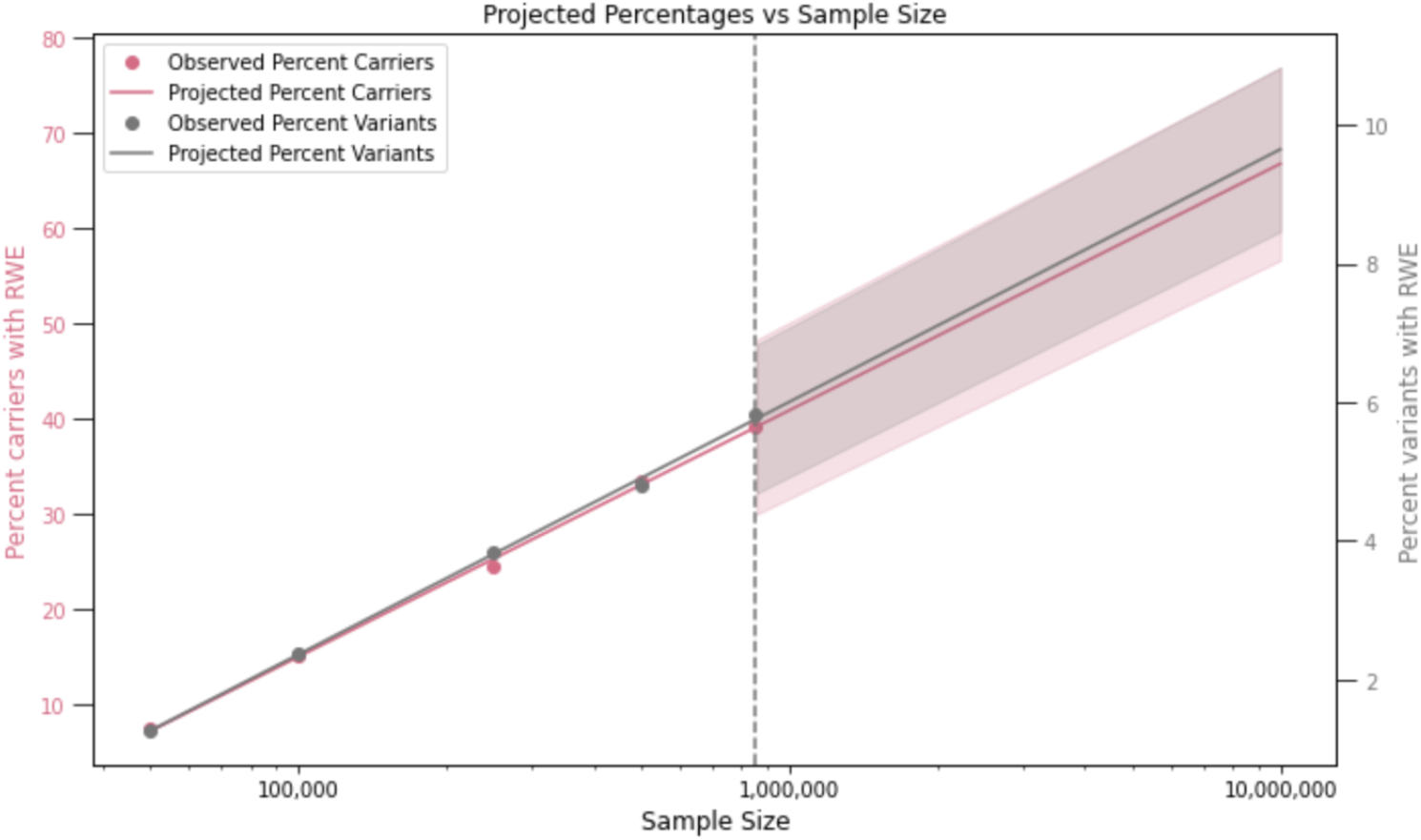
Increasing RWE sample size increases percent of cohort with evidence to reclassify VUS. Shown are the percent of VUS (gray) and VUS carriers (pink) with RWE at increasing sample sizes. The dotted line indicates the sample size used in the present study. Numbers are projected out past this sample size based on the trend, with 95%CIs.

### Limitations and Future directions

While this work showcases the potential of adding RWE to an interpretation through calibration with likelihood ratios, community-supported decisions on how best to combine this information with the other various evidence categories are needed to fully integrate this data source into current interpretation practices. Statistical thresholds for each level of evidence can also be further calibrated on a gene and disease basis. For example, future versions could account for risk alleles, which often have lower penetrance than pathogenic variants and may require different statistical thresholds. In addition, large clinicogenomic datasets of adult participants can also be leveraged as a false positive filter for highly penetrant pediatric rare disease assessments. Finally, while this manuscript focused on classically defined phenotypes for the conditions considered, we find that methods in development by Helix to utilize more sophisticated phenotypes, for example to identify more nuanced information from individuals’ medical records using large language models (LLMs), improve power for identifying genetic associations and make for even more precise pathogenicity calls.

## Conclusion

Longitudinal healthcare data, when combined with exome or genome-level sequencing, has the potential to help resolve the current VUS overload plaguing diagnostic variant interpretation and as a result improve patient care.

## Data Availability

UKB data are available for download (https://www.ukbiobank.ac.uk/) to qualified researchers. The HRN data are available to qualified researchers upon reasonable request and with permission of the HRN Steering Committee and Helix. Researchers who would like to obtain the raw genotype data related to this study will be presented with a Data Use Agreement which requires that participants will not be reidentified and no data will be shared between individuals, third parties, or uploaded onto public domains. The HRN encourages collaboration with scientific researchers on an individual basis. Examples of restrictions that will be considered in requests to data access include but are not limited to: 1. Whether the request comes from an academic institution in good standing and will collaborate with our team to protect the privacy of the participants and the security of the data requested 2. Type and amount of data requested 3. Feasibility of the research suggested 4. Amount of resource allocation for Helix and HRN member institutions required to support a collaboration.

## Acknowledgements

Funding was provided to the Healthy Nevada Project by the Renown Institute for Health Innovation and the Renown Health Foundation. The Healthy Nevada Project receives funding from Gilead Sciences, outside the scope of this research. Funding was provided to the myGenetics program by HealthPartners. We acknowledge the entire Helix bioinformatics and lab teams for their contributions to the production of exome sequencing pipeline as well as the research administration team for coordinating the project. We thank all of the participants of the Helix Research Network. This research has been conducted using the UK Biobank resource under application number 40436. This work uses data provided by patients and collected by the NHS as part of their care and support, re-used with the permission of the UK Biobank under Application Number 40436. Copyright © (2023), NHS England. All rights reserved. This research also used data assets made available by National Safe Haven as part of the Data and Connectivity National Core Study, led by Health Data Research UK in partnership with the Office for National Statistics and funded by UK Research and Innovation(grant ref MC_PC_20058).

We gratefully acknowledge All of Us participants for their contributions, without whom this research would not have been possible. We also thank the National Institutes of Health’s All of Us Research Program for making available the participant data examined in this study.

