## Supplementary Tables and Figures for "From Uncertain to Actionable: Significant Reduction in Variants of Uncertain Significance in Hereditary Germline Testing via Multi-Institutional Real-World Evidence"

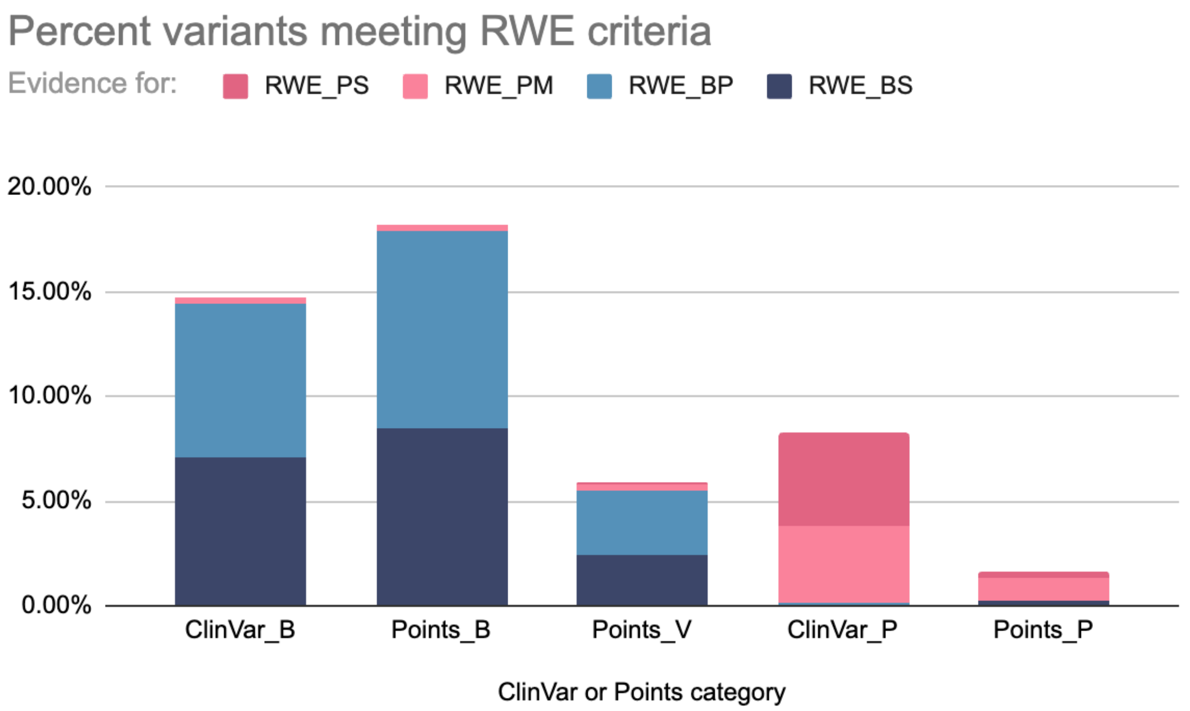


**Figure S1. Percent of variants meeting RWE criteria.** Split by established pathogenicity category from ClinVar or via automated ACMG/AMP scoring.

**Table S1. RWE criteria.**

| | Evidence for RWE_PS | | | Evidence for RWE_PM | | | Evidence for RWE_BM | | | Evidence for RWE_BS | | |
|---|---|---|---|---|---|---|---|---|---|---|---|---|
| Phenotype | Min Effect | Min 95%CI Effect | Min n case carriers | Min Effect | Min 95%CI Effect | Min n case carriers | Max Effect | Max 95%CI Effect | Min n carriers | Max Effect | Max 95%CI Effect | Min n carriers |
| Breast or Ovarian Cancer* | 4 | 2 | 5 | 4 | 2 | 3 | 2 | 4 | 8 | 1.5 | 2 | 20 |
| Breast Cancer | 2 | 1.5 | 5 | 2 | 1.5 | 3 | 1.5 | 2 | 15 | 1.25 | 1.5 | 38 |
| Colon or Endometrial Cancer | 6 | 2 | 5 | 4 | 2 | 3 | 2 | 4 | 32 | 1.5 | 4 | 79 |
| LDL levels** | 0.5 | 0.5 | 5 | 0.5 | 0.25 | 3 | 0.25 | 0.5 | 3 | 0.25 | 0.25 | 5 |

* for *BRCA1* and *BRCA2*; **including statin use as a covariate

**Table S2. All analyzed variants and their stats.** Available for download at https://www.helix.com/events/table-s2-download.

**Table S3. Alignment between RWE and ClinVar Pathogenic and Benign calls (automated ACMG/AMP scoring excluded)**

| RWE Category | Established P/LP | VUS | Established B/LB | LR toward Pathogenicity |
|---|---|---|---|---|
| RWE_BS | 1 (0.05%) | 696 (2.79%) | 552 (7.08%) | 0.01 |
| RWE_BM | 2 (0.09%) | 853 (3.43%) | 571 (7.33%) | 0.01 |
| None | 1948 (91.76%) | 23256 (93.38%) | 6646 (85.27%) | 1.08 |
| RWE_PM | 79 (3.72%) | 74 (0.3%) | 22 (0.28%) | 13.18 |
| RWE_PS | 93 (4.38%) | 25 (0.1%) | 3 (0.04%) | 113.81 |

*VUS excluded from LR calculations
